# Availability, scope, and quality of monkeypox clinical management guidelines globally: a systematic review

**DOI:** 10.1101/2022.06.09.22276219

**Authors:** Eika Webb, Ishmeala Rigby, Melina Michelen, Andrew Dagens, Vincent Cheng, Amanda Rojek, Dania Dahmash, Susan Khader, Keerti Gedela, Alice Norton, Muge Cevik, Erhui Cai, Eli Harriss, Samuel Lipworth, Robert Nartowski, Helen Groves, Peter Hart, Lucille Blumberg, Tom Fletcher, Shevin T Jacob, Louise Sigfrid, Peter Horby

**Author notes:** Joint first authors (Eika Webb, Ishmeala Rigby). Joint senior authors (Louise Sigfrid, Peter Horby).

## Abstract

**Background:** Monkeypox (MPX) is an important human orthopoxvirus infection. There has been an increase in MPX cases and outbreaks in endemic and non-endemic regions in recent decades. We appraised the availability, scope, quality, and inclusivity of clinical management guidelines for MPX globally.

**Methods:** For this systematic review, we searched six databases from inception until 14 Oct. 2021, augmented by a grey literature search until 17 May 2022. MPX guidelines providing treatment and supportive care recommendations were included, with no exclusions for language. Two reviewers assessed the guidelines. Quality was assessed using the Appraisal of Guidelines for Research and Evaluation (AGREE) II tool.

**Results:** Of 2026 records screened, 14 guidelines were included. Overall, most guidelines were of low-quality with a median score of 2 out of 7 (range: 1-7), lacked detail and covered a narrow range of topics. Most guidelines focused on adults, five (36%) provided some advice for children, three (21%) for pregnant women, and three (21%) for people living with HIV. Treatment guidance was mostly limited to advise on antivirals; seven guidelines advised cidofovir (four specified for severe MPX only); 29% (4/14) tecovirimat, and 7% (1/14) brincidofovir. Only one guideline provided recommendations on supportive care and treatment of complications. All guidelines recommended vaccination as post-exposure prophylaxis (PEP). Three guidelines advised on vaccinia immune globulin as PEP for severe cases in people with immunosuppression.

**Conclusion:** Our results highlight a concerning lack of evidence-based clinical management guidelines for MPX globally. There is a clear and urgent need for research into treatment and prophylaxis including for different risk populations. The current outbreak provides an opportunity to accelerate this research through coordinated high-quality studies. New evidence should be incorporated into globally accessible guidelines, to benefit patient and epidemic outcomes. A ‘living guideline’ framework is recommended.

**Systematic review registration:** PROSPERO CRD42020167361

**Funding statement:** This work was supported by the UK Foreign, Commonwealth and Development Office and Wellcome [215091/Z/18/Z] and the Bill & Melinda Gates Foundation [OPP1209135].

## Introduction

Monkeypox (MPX) is a zoonotic disease caused by an Orthopoxvirus belonging to the same genus as smallpox. The MPX virus was discovered in 1958,^1–4^ with the first human infection identified in 1970 in the Democratic Republic of Congo (DRC).^5^ Since then human MPX has mostly been reported in Central and Western African countries. Two distinct genetic clades of the virus have been identified – the Congo Basin and the West African clades, with a case fatality rate of 1-3.6% and 10.6%, respectively estimated in earlier outbreaks.^6^ The number of human monkeypox cases has been rising since the 1970s, with the highest increases reported in the DRC and an increase in travel-imported cases outside of Africa. In 2003, 37 confirmed cases were detected in the US, linked to contact with pet prairie dogs infected by rodents from Africa.^7,8^ This was followed by sporadic travel imported cases in the UK (2018, 2021), Israel (2018), Singapore (2019).^9–12^ From December 2021 to 1 May 2022 there were 1315 cases and 57 deaths reported from four countries in Africa.^6^

The ongoing outbreak in 2022 is the first documented multi-country outbreaks in non-endemic countries, with 257 confirmed cases in 23 countries reported as of 26^th^ May 2022.^13, 14^ The current outbreaks are assessed by the WHO as medium risk for the general population with low risk for pandemic potential. MPX presents as a vesicular-pustular illness, which may be preceded by fever, headache, tonsillitis, cough, myalgia and fatigue.^15^ Fever can be absent. Lymphadenopathy if present may distinguish it from chickenpox and smallpox.^16,17^ Complications include painful lesions, secondary infections, bronchopneumonia, encephalitis, keratitis and psychological symptoms.^15–18^ Younger children and pregnant women are at higher risk of severe disease.^15^ The incubation period is up to 21 days. Interactions with infected animals and individuals is associated with risk of infection.^19^ Human-to-human transmission occurs through direct contact (body fluids, skin lesions, mucosal surfaces, respiratory droplets), indirectly (contaminated objects), and vertically from mother-to-foetus through the placenta.^18,20,21^ Polymerase chain reaction (PCR) is the preferred diagnostic test. ^22^ Due to orthopoxviruses serological cross-reactivity, antigen and antibody detection methods do not provide MPX-specific confirmation. Previous smallpox vaccination may lead to false positive results.^18^ The smallpox vaccine has been estimated to be 85% protective against MPX.^23,24^ The first generation live smallpox vaccine is not recommended in pregnancy or in people with immunosuppression.^25,26^ Newer third generation live, non-replicating vaccines, are approved in certain regions for smallpox and monkeypox in adults.^27^ None are part of routine vaccination programs, and not readily available for public use globally.^28^

Therapeutic options are limited. Tecovirimat is licensed in some countries for the treatment of smallpox in adults and children (>13kg),^29^ and MPX during outbreaks. ^17^ Two other treatments; cidofovir and brincidofovir have been shown to be active against poxviruses,^30–32^ with cidofovir having broad-spectrum activity against DNA viruses, including herpes-, adeno-, polyoma-, papilloma-and poxviruses.^31^ Both have been shown efficacy in *in vitro* and animal studies but data on treatment in humans with MPX is limited,^32^ and they are only authorised for use in certain countries.

Even when the evidence-base is limited, clinical management guidelines are important tools for guiding clinical decision-making, and standardising the best available care between sites.^33–35^ Guidelines must be readily available, of good quality and inclusive of vulnerable patient groups. Standardisation of care will benefit patients and can also facilitate the implementation of needed multi-sites trials for therapeutics and vaccines. The increase in MPX cases in recent decades highlights the need to ensure that clinicians worldwide have access to clinical management guidelines to guide treatment, to benefit patient care and outcomes. This review aims to assess the availability, quality, scope, and inclusivity of clinical guidelines for MPX.

## Methods

This is a systematic review of the availability, inclusivity, scope, and quality of clinical management guidelines for MPX.^36^ We included guidelines that provided advice on treatment or supportive care for MPX.^37^ This study is nested within an extensive systematic review of supportive care and clinical management guidelines for high consequence infectious diseases. The study is registered with the prospective international register of systematic reviews (PROSPERO) (CRD42020167361)^38^ and follows the Preferred Reporting Items for Systematic Reviews and Meta-Analyses (PRISMA) guidelines on the conduct of systematic reviews.

### Search strategy

We searched Ovid Medline, Ovid Embase, Ovid Global Health, Scopus, Web of Science Core Collection, and WHO Global Index Medicus from inception until 14^th^ October 2021,using predefined MESH words (supplement).supplemental file 1). Search strategies applied the Canadian Agency for Drugs and Technologies in Health (CADTH) database guideline search filter.^39^ No limits were applied to the search results. We augmented this with an extensive grey literature search using Google and Google Scholar, until 17 May 2022, in Arabic, English, French, German, Mandarin, Russian and Spanish. The full search strategy is shown in the supplementary material.

### Eligibility criteria

Guidelines that included advice on treatment and supportive care for MPX were included. Guidelines that purely focused on public health or diagnostics were excluded if they did not provide any treatment advice. Local hospital standard operation protocols were excluded, we made no exclusions on languages.

### Screening and data extraction

Two reviewers independently screened the guidelines for inclusion and extracted data using Rayyan systematic review software.^40^ Data were extracted using a standardised form, previously piloted for related reviews.^41,42^ For each guideline data on source, target population, and clinical topics were extracted (Supplementary file 2). Disagreements were resolved via consensus or by a third reviewer. For non-English guidelines, team members with good to excellent knowledge of the language assessed the guidelines.

### Quality appraisal

Two reviewers independently appraised the quality using the Appraisal of Guidelines for Research and Evaluation II (AGREE II) Instrument.^43^ The AGREE II tool provides an objective framework which assesses the guideline quality based on the development process, it does not assess the validity of recommendations. The tool consists of six domains and two global ratings. The six domains are: scope and purpose, stakeholder involvement, rigour of development, clarity of presentation, applicability, and editorial independence. The score was completed by two independent assessors. There are several sub-criteria within each domain which are scored based on whether the criteria are met using a seven-point Likert scale, from 1 (strongly disagree) to 7 (strongly agree).^43^ A score of one is given when there is no information relevant to the AGREE II item provided. Guidelines were assessed as of high-quality if they scored more than 60% in at least three domains including domain three (rigour of development), as this is considered a high-quality indicator. They were assessed as of moderate quality if they scored more than 60% in at least three domains, but not in domain three, and low if they did not reach any of these criteria.^43^ Graphics were produced using R version 4.0.2.

### Patient and public involvement

There was no public or patient involvement in the course of this project due to the pandemic constraints.

## Results

Of the 2026 records screened, 14 guidelines met the eligibility criteria for inclusion.^18,44–56^ (Figure 1) Forty-three percent (6/14) were aimed for global use, 21% (3/14) for Asia, 21% (3/14) for Europe, 7% (1/14) for Africa and 7% (1/14) for North America (Table 1). Most were produced by organisations in high-or upper-middle income countries. 86% (12/14) were in English, 14% (2/14) in Mandarin.^45,54^ There was a lack of comprehensive clinical management guidelines identified, only one guideline, which was produced by the Nigerian Centre for Disease Control (NCDC) provided more detailed guidance including detailed recommendations on supportive care and treatment of complications.^44^ The guidelines made limited provision for different risk groups such as children, pregnant women and people living with HIV or immunocompromised patients.

**Table 1.** Characteristics of the included guidelines. A: Adults, C: Children, P: Pregnant Women O: Older adults, H: People living with HIV/Immunosuppression

| Guideline | Country (Region) | Year | Country income classification* | Target populations | Overall quality score |
| --- | --- | --- | --- | --- | --- |
| China (MoH)<sup>54</sup> | China | 2003 | UMC | A | 1 |
| Dermatology Advisor<sup>47</sup> | Global | 2017 | - | C, P, A, O, H | 1 |
| Dermnet<sup>48</sup> | Global | 2014 | - | C, A | 2 |
| ECDC<sup>56</sup> | Europe | 2019 | - | A, H | 1 |
| Emedicine<sup>49</sup> | Global | 2020 | - | A | 1 |
| Ireland HPSC<sup>53</sup> | Ireland | 2021 | HIC | A | 1 |
| Medscape<sup>51</sup> | Global | 2019 | - | A | 2 |
| NCDC<sup>44</sup> | Nigeria | 2019 | LMIC | C, A, H | 3 |
| PHE/UKHSA<sup>52</sup> | England | 2019 | HIC | A | 1 |
| Singapore FETP<sup>46</sup> | Singapore | 2020 | HIC | A | 3 |
| Taiwan CDC<sup>45</sup> | Taiwan | 2009 | UMC | C, P, A, H | 1 |
| Uptodate<sup>50</sup> | Global | 2021 | - | C, P, A | 6 |
| US CDC<sup>55</sup> | USA | 2018 | HIC | A | 2 |
| WHO<sup>18</sup> | Global | 2019 | - | A | 1 |

**Abbreviations:** CDC: Centers for Disease Control, ECDC: European Centres for Disease Control and Prevention, FETP: Singapore Field Epidemiology Training Program, HIC: High-income country, HPSC: Health Protection Surveillance Centre, LMIC: Lower-middle income country, MoH: Ministry of Health, NCDC: Nigeria Center for Disease Control, PHE: Public Health England, UKHSA: United Kingdom Health Security Agency, UMC: Upper-Middle Income Country, US: United State of America, WHO: World Health Organisation.<sup>57</sup>

**Figure 1.**
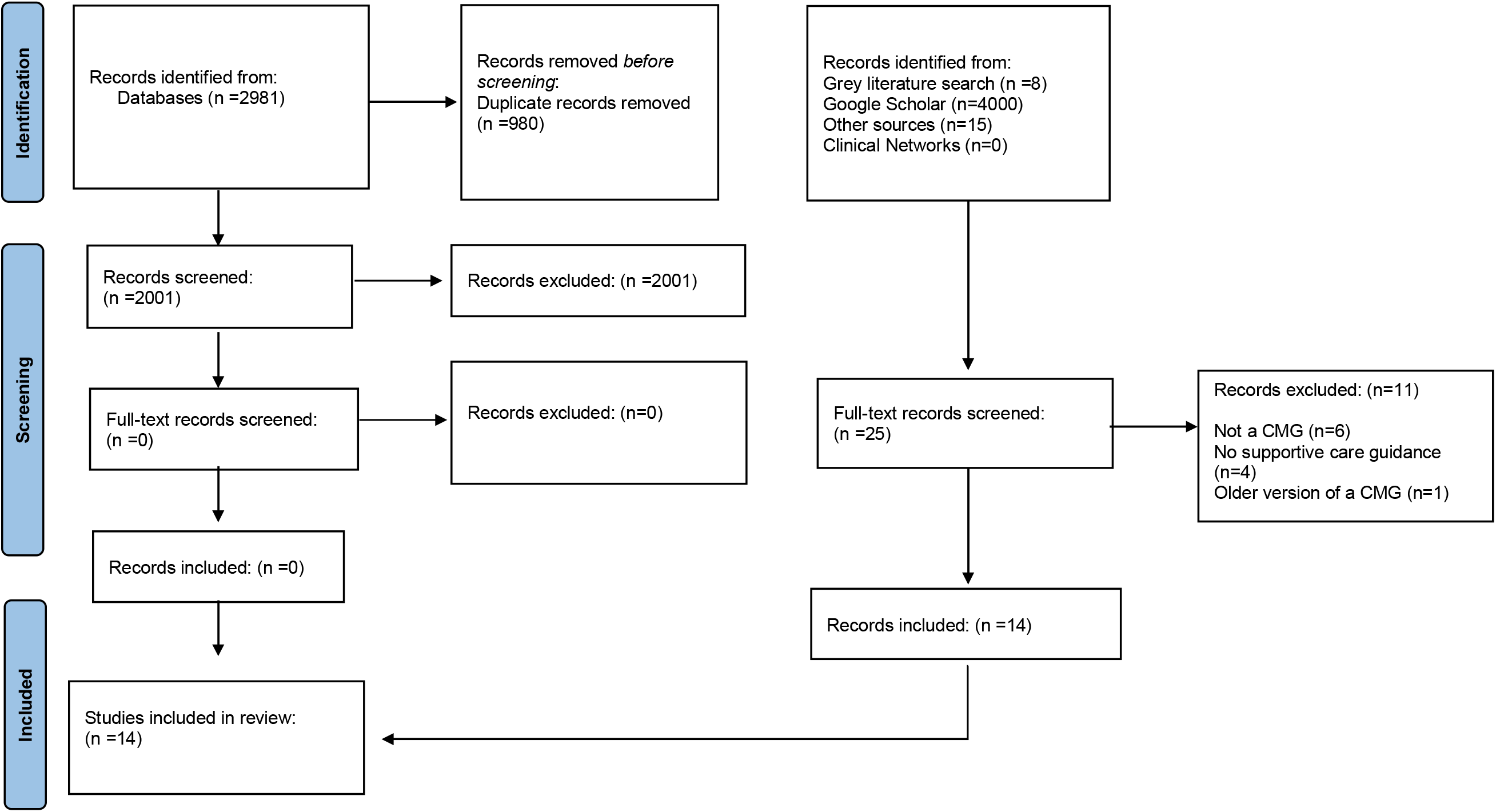
PRISMA Diagram.

### Quality assessment

Overall quality was low (Figure 2).^43^ The median overall quality was 2 out of 7 points (range: 1-7). Only one guideline was assessed as of high quality.^50^ The domain that scored the highest across the guidelines was clarity of presentation (median (IQR): 61% (50-64)). Domains in which all of the guidelines scored poorly in were rigour of development (median (IQR): 16% (8-20)), applicability (median (IQR): 15% (12-21)), scope and purpose (median (IQR): 19% (19-42)), stakeholder involvement (median (IQR): 22% (19-44)) and editorial independence 0% (0-33). The low score for certain domains, such as editorial independence, may be partly due to a lack of information provided. We observed a lack of documentation of the methodology used to develop the guidance, few guidelines used systematic reviews, and clear links to evidence were lacking. Often clinical guidance was embedded within a document that primarily focused on infection control.

**Figure 2.**
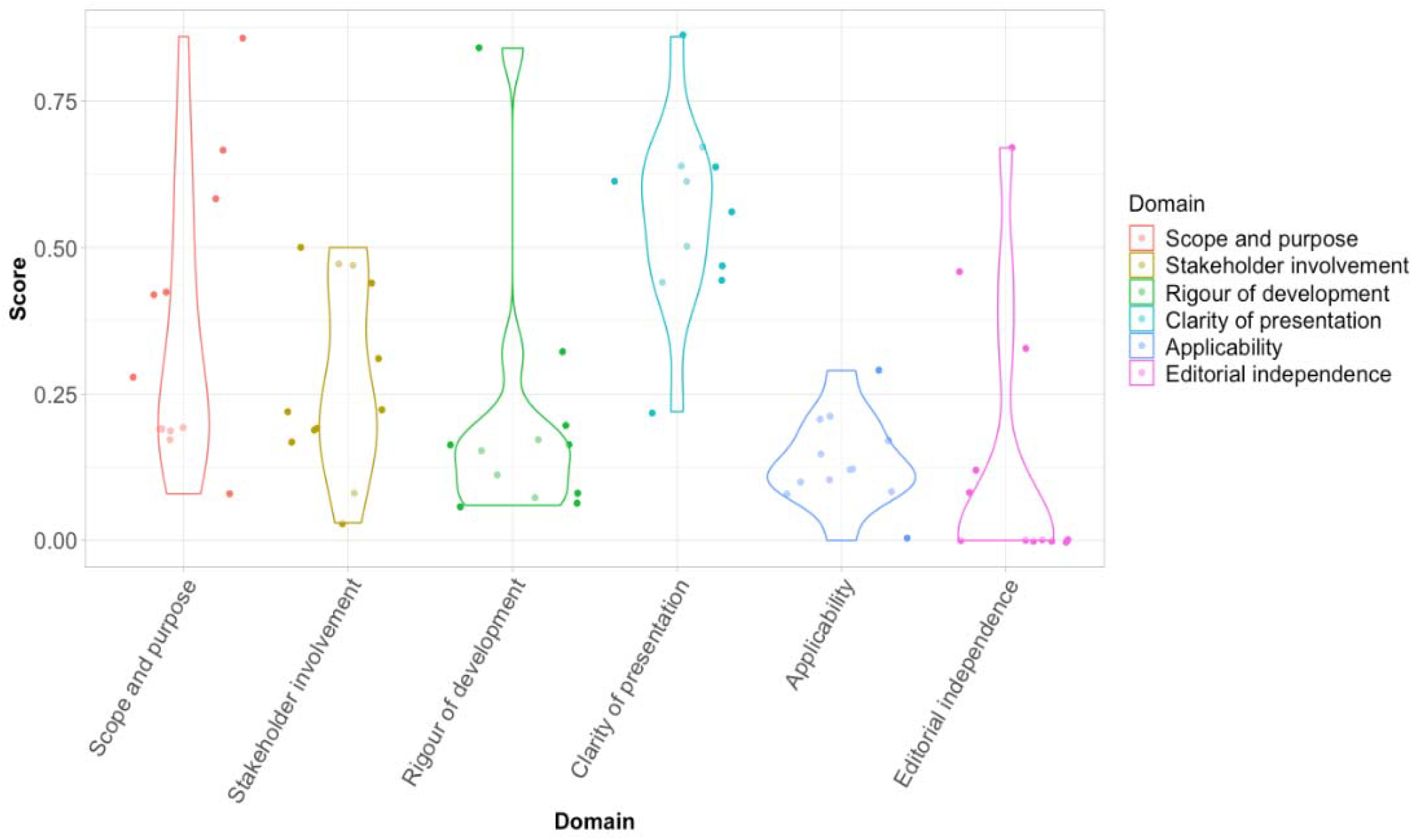
Combined AGREE II assessment of the guidelines. Figure legend: The violin plots depict the variation in scores of individual CMGs in each domain. Each dot represents a CMG’s proportional score per domain. The width of each curve represents the frequency of CMGs scoring that corresponding value in each domain. Abbreviations: AGREE: Appraisal of Guidelines for Research and Evaluation II

### Treatment recommendations

Generally, the clinical recommendations provided by the guidelines were non-specific and covered a narrow range of topics (Table 2).

**Table 2.** Overview of the recommendations provided in the guidelines

| Guideline | Country | Year | Symptom management | Antivirals | Antibiotics | Prophylaxis |
| --- | --- | --- | --- | --- | --- | --- |
| China (MoH)<sup>54</sup> | China | 2003 | R | NS | R* | R |
| Dermatology Advisor<sup>47</sup> | Global | 2017 | NS | C | NS | R |
| Dermnet<sup>48</sup> | Global | 2014 | NS | C | NS | R |
| ECDC<sup>56</sup> | Europe | 2019 | NS | NS | NS | R |
| Emedicine<sup>49</sup> | Global | 2020 | NS | C | NS | R |
| Ireland HPSC<sup>53</sup> | Ireland | 2021 | NS | C | NS | R |
| Medscape<sup>51</sup> | Global | 2019 | R | NS | NS | R |
| NCDC<sup>44</sup> | Nigeria | 2019 | R | NS | R* | R |
| PHE/UKHSA<sup>52</sup> | England | 2019 | NS | C | NS | R |
| Singapore FETP<sup>46</sup> | Singapore | 2020 | NS | NS | NS | R |
| Taiwan CDC<sup>45</sup> | Taiwan | 2009 | NS | NS | NS | R |
| Uptodate<sup>50</sup> | Global | 2021 | R | C | NS | R |
| US CDC<sup>55</sup> | USA | 2018 | NS | C | NS | R |
| WHO<sup>18</sup> | Global | 2019 | NS | C | NS | R |

**Abbreviations:** CDC: Centers for Disease Control, ECDC: European Centre for Disease Control and Prevention, FETP: Singapore Field Epidemiology Training Program, HPSC: Health Protection Surveillance Centre, MoH: Ministry of Health, NCDC: Nigeria Center for Disease Control, PHE: Public Health England, UKHSA: United Kingdom Health Security Agency, US: United States of America WHO: World Health Organisation
\*If secondary complications C: Considered, NS: Not specified, R: Recommended

Guidance varied, such as in recommendations on the type of antiviral drugs to consider, and type of vaccine for prophylaxis. Seven guidelines^47–52,55^ advised cidofovir, with four noting that it should only be considered in people presenting with severe illness. (Table 3) One guideline advised the use of brincidofovir as an alternative, citing an improved safety profile over cidofovir.^55^ Three guidelines advised tecovirimat^18^ or cidofovir ^50,55^ whereas a more recent guideline produced by the World Health Organisation (WHO) only advised tecovirimat as part of a clinical research study.^18^ None of the guidelines provided further details to guide optimal timing of treatment, dosage and duration. Two guidelines advised that vaccina immune globulin (VIG) may be considered in severe cases.^47,55^

**Table 3.**
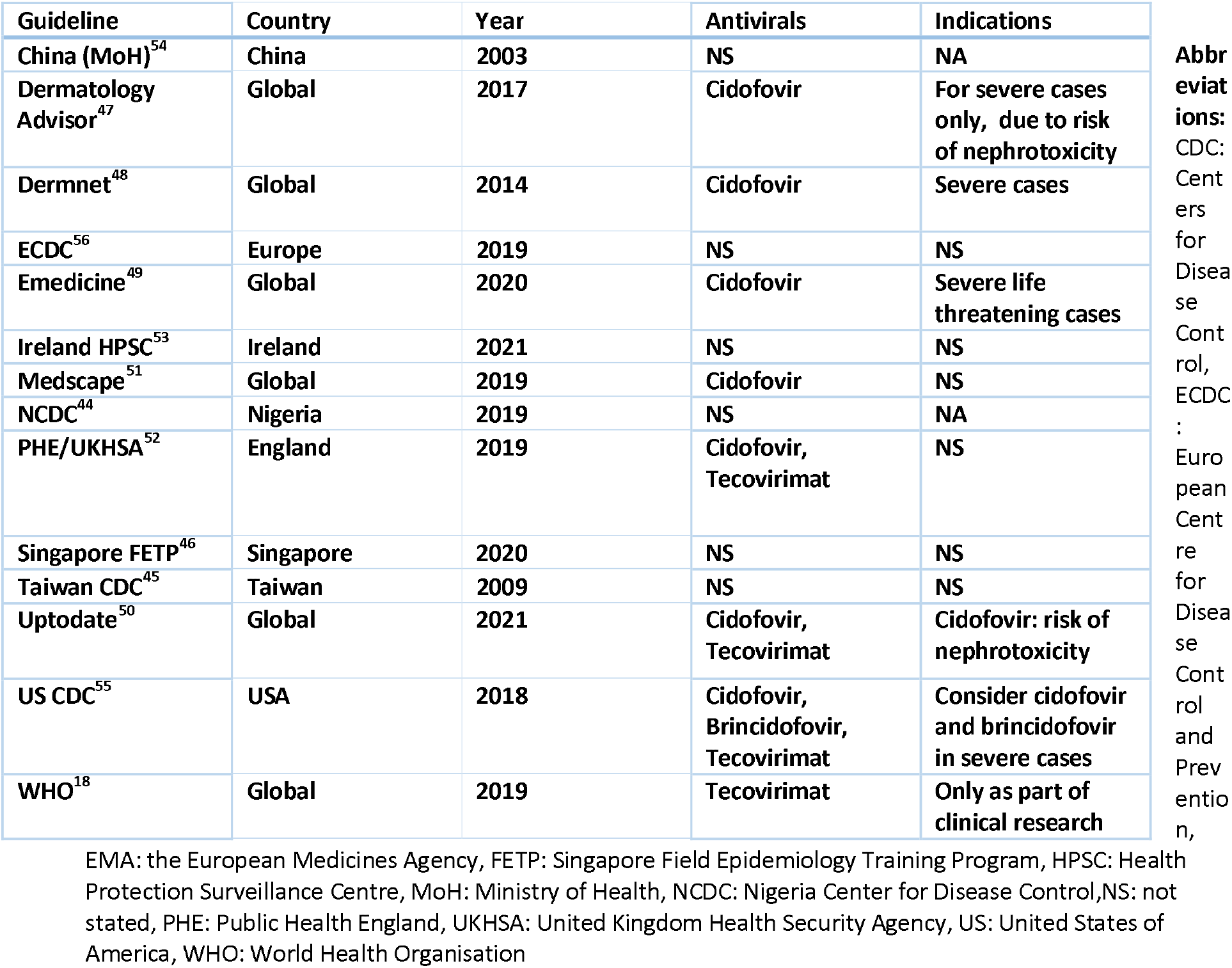
Recommendations on use of antivirals. An overview of the antiviral treatments recommended to consider in MPX. None of the guidelines provided further indications to guide optimal timing, dose, or duration of treatment.

Two guidelines recommended the use of antibiotics for the treatment of secondary complications. (Table 4).^44,54^ The guideline produced by the NCDC was the only one providing detailed recommendations on supportive care and treatment of complications, such as secondary infections and sepsis, bronchopneumonia, encephalitis, ophthalmology and psychological complications.^44^ Empirical oral or parenteral cephalosporins or beta-lactam antibiotics were recommended for the treatment of secondary bacterial infections (e.g., boils, abscesses, skin dermatitis). Empirical broad-spectrum antibiotics were advised for bronchopneumonia and encephalitis.^54^ In patients with encephalitis, they further advised close monitoring of nutrition/hydration, and consideration of nasogastric (NG) feeding for unconscious patients and anticonvulsants for seizure control.^44^ Supportive care recommendations covered the management of rashes, pruritis and ulcers (antiseptic cleaning, saline baths, antihistamines); antipyretics (paracetamol, NSAID) to manage fever and pain, and metoclopramide (IV) for adults and chlorphenamine syrup for children for nausea and vomiting. For dehydration, they advised using oral rehydration salts, particularly in children, and intravenous fluids (0.9% saline or dextrose) as indicated. Only one additional guideline provided advise on the monitoring of fluid balance, advising that patients experiencing nausea, vomiting, or dysphagia may require hospital admission for intravenous hydration.^50^

**Table 4.** Recommendations on use of antibiotics and immunoglobulins.

| Guideline | Country | Year | Antibiotics | Indications | Immunoglobulins | Indications |
| --- | --- | --- | --- | --- | --- | --- |
| China (MoH)<sup>54</sup> | China | 2003 | R | Secondary bacterial infections | NS | - |
| Dermatology Advisor<sup>47</sup> | Global | 2017 | NS | - | VIG | Consider in severe infection |
| Dermnet<sup>48</sup> | Global | 2014 | NS | - | NS | - |
| ECDC<sup>56</sup> | Europe | 2019 | NS | - | NS | - |
| Emedicine<sup>49</sup> | Global | 2020 | NS | - | NS | Notes that VIG has not shown efficacy for treatment |
| Ireland HPSC<sup>53</sup> | Ireland | 2021 | NS | - | NS | - |
| Medscape<sup>51</sup> | Global | 2019 | NS | - | NS | Notes that VIG has not shown efficacy in treatment |
| NCDC<sup>44</sup> | Nigeria | 2019 | R | Secondary bacterial infections<br>Cefuroxime 500mg bd, 5days (oral/parental) or<br>Ceftriaxone IV 1g , 5 days or<br>B-lactam antibiotics | NS | - |

|  |  |  |  | (Amoxyl/Clavulanic acid, 625mg x2/day, ≥ 5days) |  |  |
| --- | --- | --- | --- | --- | --- | --- |
| PHE/UKHSA<sup>52</sup> | England | 2019 | NS | - | NS | - |
| Singapore FETP<sup>46</sup> | Singapore | 2020 | NS | - | NS | - |
| Taiwan CDC<sup>45</sup> | Taiwan | 2009 | NS | - | NS | - |
| Uptodate<sup>50</sup> | Global | 2021 | NS | - | NS | - |
| US CDC<sup>55</sup> | USA | 2018 | NS | - | VIG | Can be considered in severe cases |
| WHO<sup>18</sup> | Global | 2019 | NS | - | NS | - |

**Abbreviations:** CDC: Centers for Disease Control, ECDC: European Centre for Disease Control and Prevention, FETP: Singapore Field Epidemiology Training Program, HPSC: Health Protection Surveillance Centre, MoH: Ministry of Health, NA: Not applicable, NCDC: Nigeria Center for Disease Control, PHE: Public Health England, UKHSA: United Kingdom Health Security Agency, US: United States of America, VIG: Vaccinia immune globulin, WHO: World Health Organisation

### Recommendations on pre-and post-exposure prophylaxis (PEP)

The older generation smallpox vaccines is no longer part of routine immunisation programs.^18^ There have been several developments of modified smallpox vaccines in recent years, including second generation vaccines such as ACAM2000^25^ which was recommended for PEP in three guidelines (Table 5).^44,49,51^ A third-generation vaccine, commonly known as Imvamune/Imvanex or Jynneos was recommended for PEP by seven guidelines. Only two guidelines provided advice on the optimal timing of PEP.^46,49^ The guidance on PEP for different at risk populations were limited and at times conflicting. Two guidelines provided advised on PEP in children and pregnant women,^50^ one stating that although smallpox vaccination may be contraindicated by pregnancy, age, and a history of eczema in the pre-event context, they can be used with caution in the event of exposure.^47^ Another guideline advised against vaccination of infants (< 1 years old) and pregnant women.^45^ Two guidelines specifically recommended against the use of the vaccinia smallpox vaccine in people with immunosuppression (i.e., in people with HIV and a CD4 counts<200, or on chemotherapy).^45,47^ The guidance on the use of VIG was contradictory. Three guidelines advised considering VIG in individuals with compromised immune function^47,50,55^ whereas two guidelines did not provide any recommendations on its use, but advised that data on its effectiveness for treatment and PEP is lacking.^49,51^ Six guidelines recommended immunisation of people at risk of MPX exposure such as healthcare workers.^48,49,51,53–55^

**Table 5.** Recommandations on pre-and post-exposure prophylaxis.

| Guideline | Country | Year | Vaccination | Indications | Immunoglobulins | Indications |
| --- | --- | --- | --- | --- | --- | --- |
| China (MoH)<sup>54</sup> | China | 2003 | A smallpox vaccine | Individuals after contact with suspected animal or human case. | NS | - |
| Dermatology Advisor<sup>47</sup> | Global | 2017 | A smallpox vaccine | Individuals at risk of infection prior to exposure.<br>Contraindication: immune-compromised individuals i.e. T-cell deficiency, HIV with CD4<200, or by medication. | VIG | In immune-compromised patients. Notes that VIG should only be used in severe disease |
| Dermnet<sup>48</sup> | Global | 2014 | A smallpox vaccine | All healthcare workers and all close contacts with infected cases. | NS | - |
| ECDC<sup>56</sup> | Europe | 2019 | MVA-BN/Imvanex | NS | NS | - |
| Emedicine<sup>49</sup> | Global | 2020 | ACAM2000 and Jynneos | ≤ 2 weeks, ideally ≤ 4 days. Exposed health care workers, household contacts of confirmed cases. Note: for ACAM2000 avoid risk of spread from inoculation site to other sites and individuals | NS | Notes that VIG has not shown efficacy as prophylaxis |
| Ireland HPSC<sup>53</sup> | Ireland | 2021 | Vaccinia, Imvanex (3<sup>rd</sup> gen) | Imvanex for healthcare workers, close contacts including in outbreak settings and first responders. Can be used for individuals for whom previous smallpox vaccinations were contraindicated | NS | - |
| Medscape<sup>51</sup> | Global | 2019 | Jynneos, ACAM2000 | Exposed healthcare workers and household contacts of confirmed cases. Note: care to avoid spread from inoculation site (ACAM 2000) | NS | Notes that VIG has not shown efficacy as prophylaxis |
| NCDC<sup>44</sup> | Nigeria | 2019 | ACAM2000, Imvamune (3rd gen) | NS | NS | - |
| PHE/UKHSA<sup>52</sup> | England | 2019 | A smallpox vaccine | NS | NS | - |
| Singapore FETP<sup>46</sup> | Singapore | 2020 | A smallpox vaccine | Post-exposure prophylaxis within 4 days, up to 14 days | NS | - |
| Taiwan CDC<sup>45</sup> | Taiwan | 2009 | Vaccinia | People who care for sick patients, or animals, study the virus, or MPX epidemics who have not been vaccinated, should be vaccinated. | NS | - |
| Uptodate<sup>50</sup> | Global | 2021 | MVA, Imvamune and Jynneos) | Contacts (expect immunocompromised patients) | VIG | if immunocompromised |
| US CDC<sup>55</sup> | USA | 2018 | Jynneos (Imvamune, Imvanex) | For contacts of cases | VIG | If severe immunodeficiency, T-cell dysfunction if smallpox vaccination is contraindicated |
| WHO<sup>18</sup> | Global | 2019 | A smallpox vaccine | NS | NS | - |

**Abbreviations:** CDC: Centers for Disease Control, ECDC: European Centre for Disease Control and Prevention, FETP: Singapore Field Epidemiology Training Program, HPSC: Health Protection Surveillance Centre, MoH: Ministry of Health, MVA- BN: Modified Vaccinia Ankara - Bavarian Nordic, NCDC: Nigeria Center for Disease Control, NS: Not Stated, PHE: Public Health England, UKHSA: United Kingdom Health Security Agency, US: United States of America, VIG: Vaccinia Immune Globulin, WHO: World Health Organisation

### Infection prevention measures

Most guidelines (n=13) provided some advice on infection prevention measures in healthcare settings.^18,44–47,49–56^ Eight guidelines advised on the isolation of patients with suspected MPX infection.^44–46,51–55^ One advised isolation until all lesions are crusted and dry,^44^ another till all crusts have fallen off and the skin healed.^51^ Six guidelines provided advice on eye protection for procedures with risk of body fluid exposure, and five advocated for the use of facemasks,^44,47,54,55^ of which three specified N95 masks in healthcare settings.^44,45,54^

## Discussion

Our review identified a lack of up-to-date, high quality evidence-based clinical management guidelines for MPX infection. As we continue to experience an increase in MPX cases and outbreaks including in regions, with limited clinical experience in managing cases, there is a need for clinical management guidelines to guide patient care. Clinical management guidelines are important tools for frontline clinicians during outbreaks. Guidelines should be developed using robust methodologies for clinicians to be able to assess their validity. However, we found that most guidelines did not document the methodology used, which is reflected in the quality assessment, with most guidelines identified assessed as of poor quality. The low scores seen for the rigour of development reflect a lack of systematic methods, documentation and clear links to the evidence supporting recommendations.

The most marked difference across the guidelines was the antivirals and vaccines recommendations. Most guidelines that advised antiviral treatments recommended cidofovir, whereas more recently updated guidelines, such as the WHO guideline advised to consider tecovirimat.^18^ Similar variations in guidance, was observed for post-exposure prophylaxis, with more recently updated guidelines advising use of the newer generation smallpox/MPX vaccines.^44,49–51,53,55,56^ This highlights a fundamental issue in the development of guidelines for the management of neglected infectious diseases, which was also observed in other reviews ^42,58^ We observe a tendency of guidelines being developed rapidly in response to outbreaks, never to be revisited again, but still being available in public domains. Failure to recall out of date guidelines as new evidence emerges, pose a risk to patient care. Few guidelines report mechanisms for updates or monitoring.

Our review also identified a concerning lack of guidance on the treatment and PEP, and at times contradictory advice, for different population groups such as children, pregnant women and people living with immunosuppression, which could exacerbate their vulnerability in outbreaks.

Variations in the recommendations may reflect that some guidelines were produced before newer treatments were authorized in various regions. Most are only authorized in a limited number of countries, which raises important questions on equity in access to best available care worldwide. Considering MPX is a mild disease in most, there was a surprising lack of advice on the management based on the severity of illness. Only one guideline identified produced by NCDC gave detailed supportive care recommendations, including on the management of symptoms and secondary complications, such as bacterial infections, encephalitis, ophthalmological and psychological conditions.^44^ Many guidelines were positioned within public health guidance, which may partly explain the limited details provided to guide treatment and patient management. There is an argument for combining clinical management and hospital infection control advise to protect healthcare workers and reduce risk of nosocomial transmission. This requires that the guidance is supported by evidence, as the implementation of control measures may have wide direct and indirect impact on healthcare systems, especially in resource constraint settings and context.

Even with a limited evidence base, clinical management guidelines are important tools for guiding decision making and to reduce risk of inappropriate treatments. The variation seen, and the lack of recommendations for high-risk populations underline the importance of a gold standard framework for guideline development. The lack of clarity between guidelines creates uncertainty for clinicians treating patients with MPX which may impact patient care.

This review is not without its limitations. Firstly, most guidelines were published as grey literature, and although we made an extensive search in different languages, we may have missed additional guidelines available. Secondly, even though guidelines that required translations were assessed by a reviewer with good knowledge of the language, there is a risk that finer nuances may have been lost in translation. Thirdly, the AGREE II tool was primarily designed for guidelines produced in non-emergent settings, and although we are confident of its applicability to a variety of settings, it may contribute to the lower average quality scores. Finally, the quality assessment focuses on the development process, but does not assess the validity of the recommendations. Nevertheless, we identified limited availability of comprehensive clinical management guidelines for people affected by MPX, which may have impact on patient care.

Our study highlights a need for a rigorous framework for producing guidelines ahead of epidemics and a recognised platform for rapidly reviewing and updating guidance during outbreaks, as new evidence emerges. Human MPX is providing a challenge even in high-resource settings with well-resourced healthcare systems. The lack of guidelines may especially impact clinics with limited previous experience in managing patients with MPX. Given the recent global publicity surrounding MPX, this is an opportune moment for harnessing interest and investment for research into the efficacy of therapeutics and vaccines, to inform optimal treatment and prophylaxis strategies for the whole population.

Developing guidelines is resource intensive. The most comprehensive guideline identified was developed by a national public health organization (NCDC) based in an endemic country.^44^ This emphasizes the need for wide stakeholder involvement in guideline development, including experienced topic experts and affected communities. Guidelines developed by global organisations in collaboration with clinicians with experience in managing patients in different settings may be the optimal solution to improve quality, standardisation of recommendations and applicability. Ensuring that organisations that clinicians may turn to for guidance during outbreaks, such as the WHO, has the resources to provide the best possible guidelines and for these to be updated is important. A ‘living guideline’ framework for infectious disease is recommended to improve availability of up-to-date clinical management guidelines, developed using robust methodologies and inclusive of different at-risk populations. Urgent investments into research to identify optimal treatment and prophylaxis strategies are needed for the whole population to benefit patient care and outcomes.

## Data Availability

All data generated or analysed during this study are available on reasonable requests from the corresponding author.

## Data availability statement

### Ethics approval and consent to participate

Not Applicable

## Consent for publication

Not Applicable

## Author contributions

AD, SL, VC, LS, EH, SJ, LB, PH, HG, TF, EW, MM, IR informed the study protocol. EW, IR, LS led on writing the manuscript with input from all co-authors. EH, AD carried out the database search with input from MM, EW and IR. EW, MM, IR, AD, EC, SK conducted the grey literature search. EW, RJ, AD, MM screened the retrieved articles for inclusion. AD, EW, IR, RJ, MM extracted the data and completed the risk of bias analysis. LS, EW, MM, RJ, SL, IR, SK, MC, AR, AN lead on the data analysis and presentation of the results. LS, LS, MM, IR, STJ, TF, SK, AD, MC, AR, AN informed the interpretation of the results. LS, PH, TF and STJ Provided overall supervision, leadership and advice. PH, HG, STJ, TF, PH, LS, AD, and LB conceptualised the project. All authors reviewed and approved the final version of the manuscript.

### Competing interest

Peter Hart is a senior research advisor and Helen Groves is a research manager at the Wellcome Trust, which provided part of the funding for this work, but neither had a role in data collection, analysis nor interpretation of the findings. Wellcome supports a range of research funding activities including awards made to ISARIC.

### Funding statement

This work was supported by the UK Foreign, Commonwealth and Development Office and Wellcome [215091/Z/18/Z] and the Bill & Melinda Gates Foundation [OPP1209135]. For the purpose of Open Access, the author has applied a CC BY public copyright licence to any Author Accepted Manuscript version arising from this submission. SL is an MRC Clinical Research Training fellow (MR/T001151/1).

### Transparency statement

The lead authors (the manuscript’s guarantor) affirms that the manuscript is an honest, accurate, and transparent account of the study being reported; that no important aspects of the study have been omitted; and that any discrepancies from the study as originally planned and registered have been explained.

## List of abbreviations

AGREE: Appraisal of Guidelines for Research and Evaluation
A: Adults
C: Considered
CADTH: Canadian Agency for Drugs and Technologies in Health
CDC: Centre for Disease Control
C: Children
DRC: Democratic Republic of Congo
ECDC: European Centres for Disease Control and Prevention
FDA: Food and Drug Administration
FETP: Singapore Field Epidemiology Training Program
HIC: High-income country
HIV: Human immunodeficiency virus
HPSC: Health Protection Surveillance Centre
H: People living with HIV/Immunosuppression
ISARIC: International Severe Acute Respiratory and Emerging Infection Consortium
IQR: interquartile range
IV: Intravenous
LMIC: Lower-middle income country
MESH: Medical Subject Headings
MoH: Ministry of Health
MPX: Monkeypox
MPXV: Monkeypox virus
MVA-BN: Modified Vaccinia Ankara - Bavarian Nordic
NA: Not applicable
NCDC: Nigeria Centre for Disease Control
NG: nasogastric
NS: Not specified
NSAID: Non-steroidal anti-inflammatory drugs
O: Older adults
ORS: oral rehydration salts
PCR: Polymerase chain reaction
PEP: post-exposure prophylaxis
PHE: Public Health England
PRISMA: Preferred Reporting Items for Systematic Reviews and Meta-Analyses
P: Pregnant Women
PROSPERO: The International Prospective Register of Systematic Reviews
R: Recommended
UK: United Kingdom
UKHSA: United Kingdom Health Security Agency
UMC: Upper-Middle Income Country
US: United States of America
VIG: Vaccinia Immune Globulin
WHO: World Health Organisation

