## Supplementary material for "Availability, scope, and quality of monkeypox clinical management guidelines globally: a systematic review": Monkeypox Supplementary file 1.06.2022 .docx

**Supplemental files**

**Table of Contents**

**S1: search strategy.............................................................................................................................................................2**

Database search strategy..........................................................................................................................................................2-7

Google Scholar search strategy................................................................................................................................................8-9

Grey Literature search strategy................................................................................................................................................9-11

**S2: Data extraction..................................................................................................................................................................12**

Data extraction form.....................................................................................................................................................................12-15

**S3: Figures and Tables.................................................................................................................................................................14**

Figure S3.1 .........................................................................................................................................................................................14

**S1: SEARCH STRATEGY**

**S1.2: Database search strategy**

**Updated on 14/10/2021 by an Outreach Librarian at the Bodleian Health Care Libraries, University of Oxford.** This review is part of a wider project evaluating the availability, quality, and inclusivity of clinical management guidelines for the management of high consequence infectious diseases (PROSPERO CRD42020167361).

**Database: Medline (Ovid MEDLINE® Epub Ahead of Print, In-Process & Other Non-Indexed Citations, Ovid MEDLINE® Daily and Ovid MEDLINE®) 1946 to present**

Search Strategy:

--------------------------------------------------------------------------------

1	exp clinical pathway/ (7266)

2	exp clinical protocol/ (178784)

3	exp consensus/ (16567)

4	exp consensus development conference/ (12448)

5	exp consensus development conferences as topic/ (2968)

6	critical pathways/ (7266)

7	exp guideline/ (36281)

8	guidelines as topic/ (41609)

9	exp practice guideline/ (29146)

10	practice guidelines as topic/ (125684)

11	(guideline or practice guideline or consensus development conference or consensus development conference, NIH).pt. (46006)

12	(standards or guideline or guidelines).ti,kf,kw. (121468)

13	((practice or treatment* or clinical) adj guideline*).ab. (45501)

14	(position statement* or policy statement* or practice parameter* or best practice*).ti,ab,kf,kw. (38873)

15	(CPG or CPGs).ti. (6054)

16	consensus*.ti,kf,kw. (29948)

17	((critical or clinical or practice) adj2 (path or paths or pathway or pathways or protocol*)).ti,ab,kf,kw. (23047)

18	recommendat*.ti,kf,kw. (46828)

19	(care adj2 (standard or path or paths or pathway or pathways or map or maps or plan or plans)).ti,ab,kf,kw. (69406)

20	(algorithm* adj2 (screening or examination or test or tested or testing or assessment* or diagnosis or diagnoses or diagnosed or diagnosing or pharmacotherap* or therap* or treatment* or intervention*)).ti,ab,kf,kw. (19893)

21	1 or 2 or 3 or 4 or 5 or 6 or 7 or 8 or 9 or 10 or 11 or 12 or 13 or 14 or 15 or 16 or 17 or 18 or 19 or 20 (659924)

22	exp Henipavirus/ (809)

23	Henipavirus Infections/ (567)

24	(nipah or hendra).tw. (1353)

25	Monkeypox virus/ or Monkeypox/ (496)

26	(monkeypox or "monkey pox").tw. (803)

27	Chikungunya virus/ or Chikungunya Fever/ (3815)

28	Chikungunya.tw. (5982)

29	"Severe Fever with Thrombocyto* Syndrome".tw. (700)

30	SFTS.tw. (1052)

31	Plague/ (5399)

32	plague.tw. (9601)

33	"black death".tw. (284)

34	"pathogen x".tw. (51)

35	22 or 23 or 24 or 25 or 26 or 27 or 28 or 29 or 30 or 31 or 32 or 33 or 34 (20833)

36	21 and 35 (186)

37	limit 36 to yr="2020 - 2021" (30)

**Database: Embase 1974 to present**

Search Strategy:

--------------------------------------------------------------------------------

1	exp clinical pathway/ (8958)

2	exp clinical protocol/ (107994)

3	exp consensus/ (81891)

4	exp consensus development conference/ (24950)

5	exp consensus development conferences as topic/ (24950)

6	critical pathways/ (8958)

7	exp practice guideline/ (615078)

8	guidelines as topic/ (433775)

9	exp practice guideline/ (615078)

10	practice guidelines as topic/ (368202)

11	(guideline or practice guideline or consensus development conference or consensus development conference, NIH).pt. (0)

12	(standards or guideline or guidelines).ti,kw. (149823)

13	((practice or treatment* or clinical) adj guideline*).ab. (69128)

14	(position statement* or policy statement* or practice parameter* or best practice*).ti,ab,kw. (55950)

15	(CPG or CPGs).ti. (7254)

16	consensus*.ti,kw. (36376)

17	((critical or clinical or practice) adj2 (path or paths or pathway or pathways or protocol*)).ti,ab,kw. (34345)

18	recommendat*.ti,kw. (56910)

19	(care adj2 (standard or path or paths or pathway or pathways or map or maps or plan or plans)).ti,ab,kw. (121600)

20	(algorithm* adj2 (screening or examination or test or tested or testing or assessment* or diagnosis or diagnoses or diagnosed or diagnosing or pharmacotherap* or therap* or treatment* or intervention*)).ti,ab,kw. (28786)

21	1 or 2 or 3 or 4 or 5 or 6 or 7 or 8 or 9 or 10 or 11 or 12 or 13 or 14 or 15 or 16 or 17 or 18 or 19 or 20 (1018905)

22	exp henipavirus/ (1485)

23	exp Henipavirus infection/ (608)

24	(nipah or hendra).tw. (1533)

25	monkeypox/ or monkeypox virus/ (861)

26	(monkeypox or "monkey pox").tw. (896)

27	Chikungunya virus/ or chikungunya/ (6073)

28	Chikungunya.tw. (7360)

29	"Severe Fever with Thrombocyto* Syndrome".tw. (758)

30	SFTS.tw. (1258)

31	plague/ (6332)

32	plague.tw. (8453)

33	"black death".tw. (287)

34	"pathogen x".tw. (43)

35	22 or 23 or 24 or 25 or 26 or 27 or 28 or 29 or 30 or 31 or 32 or 33 or 34 (24000)

36	21 and 35 (453)

37	36 (453)

38	limit 37 to yr="2020 - 2021" (83)

**Database: Global Health <1973 to 2021 Week 39>**

Search Strategy:

--------------------------------------------------------------------------------

1	exp consensus/ (140)

2	guidelines/ (55586)

3	(standards or guideline or guidelines).ti. (13730)

4	((practice or treatment* or clinical) adj guideline*).ab. (6597)

5	(position statement* or policy statement* or practice parameter* or best practice*).ti,ab. (5865)

6	(CPG or CPGs).ti. (532)

7	consensus*.ti. (2284)

8	((critical or clinical or practice) adj2 (path or paths or pathway or pathways or protocol*)).ti,ab. (1890)

9	recommendat*.ti. (7489)

10	(care adj2 (standard or path or paths or pathway or pathways or map or maps or plan or plans)).ti,ab. (6793)

11	(algorithm* adj2 (screening or examination or test or tested or testing or assessment* or diagnosis or diagnoses or diagnosed or diagnosing or pharmacotherap* or therap* or treatment* or intervention*)).ti,ab. (2152)

12	1 or 2 or 3 or 4 or 5 or 6 or 7 or 8 or 9 or 10 or 11 (82545)

13	exp henipavirus/ (1143)

14	(nipah or hendra).tw. (1155)

15	exp monkeypox virus/ (388)

16	(monkeypox or "monkey pox").tw. (489)

17	exp chikungunya virus/ (4437)

18	Chikungunya.tw. (5218)

19	"Severe Fever with Thrombocyto* Syndrome".tw. (564)

20	SFTS.tw. (524)

21	plague/ (3334)

22	plague.tw. (7648)

23	"black death".tw. (109)

24	"pathogen x".tw. (4)

25	13 or 14 or 15 or 16 or 17 or 18 or 19 or 20 or 21 or 22 or 23 or 24 (15014)

26	12 and 25 (185)

27	26 (185)

28	limit 27 to yr="2020 - 2021" (26)

**Scopus**

( ( TITLE-ABS-KEY ( "clinical pathway*" OR "clinical protocol*" OR consensus OR guideline* OR "position statement*" OR "policy statement*" OR "practice parameter*" OR "best practice*" ) OR TITLE-ABS-KEY ( care W/2 ( standard OR path OR paths OR pathway OR pathways OR map OR maps OR plan OR plans ) ) OR TITLE-ABS-KEY ( algorithm* W/2 ( screening OR examination OR test OR tested OR testing OR assessment* OR diagnosis OR diagnoses OR diagnosed OR diagnosing ) ) OR TITLE-ABS-KEY ( algorithm* W/2 ( pharmacotherap* OR therap* OR treatment* OR intervention* ) ) OR TITLE ( standards OR recommendat* ) ) ) AND ( TITLE-ABS-KEY ( henipavirus OR nipah OR hendra OR monkeypox OR "monkey pox" OR chikungunya OR "Severe Fever with Thrombocyto* Syndrome" OR sfts OR plague OR "black death" OR "pathogen x" ) ) AND ( LIMIT-TO ( PUBYEAR , 2021 ) OR LIMIT-TO ( PUBYEAR , 2020 ) )

**Web of Science Core Collection**

#1 TOPIC: ("clinical pathway*" OR "clinical protocol*" OR consensus OR guideline* OR "position statement*" OR "policy statement*" OR "practice parameter*" OR "best practice*" OR CPG OR CPGs) OR TOPIC: ((care near/2 (standard or path or paths or pathway or pathways or map or maps or plan or plans))) OR TOPIC: ((algorithm* near/2 (screening or examination or test or tested or testing or assessment* or diagnosis or diagnoses or diagnosed or diagnosing))) OR TOPIC: ((algorithm* near/2 (pharmacotherap* or therap* or treatment* or intervention*))) OR TITLE: (standards OR recommendat*)

#2 TOPIC: (Henipavirus OR nipah or hendra or monkeypox or "monkey pox" OR Chikungunya OR "Severe Fever with Thrombocyto* Syndrome" OR SFTS OR plague OR "black death" OR "pathogen x")

#3 #2 AND #1

#4 **#1 and #2** and **2020** or **2021** (Publication Years)

**The WHO Global Index Medicus Regional Libraries** <https://pesquisa.bvsalud.org/gim/?lang=en>

(tw:("clinical path*" OR "clinical protocol*" or "critical path*" OR "critical protocol*" OR "practice path*" OR "practice protocol*" OR consensus OR guideline* OR standards OR "position statement*" OR "policy statement*" OR "practice parameter*" OR "best practice*" OR CPG OR CPGs OR "care standard" OR "care path*" OR "care map*" OR "care plan*" OR algorithm*)) AND (tw:(Henipavirus OR nipah or hendra or monkeypox or "monkey pox" OR Chikungunya OR "Severe Fever with Thrombocyto* Syndrome" OR SFTS OR plague OR "black death" OR "pathogen x"))

Limits: 2020-2021

**Database search results**

|  | **Search results (February 2020)** | **Updated Search results 14/10/2021 (2020-2021 only)** |
| --- | --- | --- |
| **Database** | **Cluster 3 results** |  |
| Ovid Medline | 157 | 30 |
| Ovid Embase | 382 | 83 |
| Ovid Global Health | 157 | 26 |
| Scopus | 725 | 131 |
| Web of Science Core Collection | 722 | 176 |
| WHO Global Index Medicus | 85 | 12 |
| TOTAL | 2228 | 458 |
| Total after deduplication | 1428 | 278 |

**Google Scholar – Screened the first 10 pages of results**

**Google Scholar search updated 18^th^ May 2022**

**Sorted by relevance:**

(guideline|consensus|standards|"clinical path*”|"clinical protocol*"|"practice path*"|" policy statement*"|"best practice*") (monkeypox)

<https://scholar.google.co.uk/scholar?q=(guideline%7Cconsensus%7Cstandards%7C%22clinical+path*%E2%80%9D%7C%22clinical+protocol*%22%7C%22practice+path*%22%7C%22+policy+statement*%22%7C%22best+practice*%22)(monkeypox)&hl=en&as_sdt=0,5>

**S1.2: Grey literature search strategy**

The search terms below were used to search for monkeypox clinical management guidelines on the ministry of health, public health, and other relevant websites. The first 50 results were examined. If a relevant guideline was found using the first search term, the other search terms were not used.

**Search Terms**

Clinical management guidelines for Monkeypox

Monkeypox clinical management guidelines

Monkeypox clinical guidance

Monkeypox guidelines

Monkeypox guidance

Strategies for management of Monkeypox

Clinical practice guidelines for management of Monkeypox

Clinical management of Monkeypox

Monkeypox clinical management

**S1.2.1: Websites of organisations searched:**

**Organisation**

Infectious Disease Society of America (IDSA)

European Respiratory Society (ERS)

European society of clinical microbiology and infectious diseases (ESCMID)

British Thoracic Society (BTS)

The National Institute for Health and Care Excellence (NICE)

World Health Organisation (WHO)

US Centre for Disease Control (CDC)

Africa Centre for Disease Control (CDC)

Nigeria Centre for Disease Control (CDC)

China Centre for Disease Control (CDC)

European Centre for Disease Control (CDC)

Medscape

Uptodate guidelines

BMJ Best Practice

Taiwan Centre for Disease Control (CDC)

World Health Organisation regional offices

**S1.2.2: Ministry of Health/Public Health countries’ websites searched:**

| **Ministry of health countries** | | | | | |
| --- | --- | --- | --- | --- | --- |
| **Latin America** | **Asia** | **Europe** | **Africa** | **Middle East** | **North Americas** |
| Argentina | Australia | France | Ghana | Saudia Arabia | Canada |
| Brazil | Japan | Italy | Nigeria | UAE |  |
| Mexico | South Korea | Germany | Cameroon | Egypt |  |
| Jamaica | China Taiwan | Russia | South Africa | Jordan |  |
| Cuba | China | Turkey |  | Lebanon |  |
| and other countries in the region | Malaysia | Spain |  | Tunisia |  |
|  | Thailand |  |  |  |  |
|  | Iran |  |  |  |  |
|  | India |  |  |  |  |
|  | Indonesia |  |  |  |  |
|  | Nepal |  |  |  |  |
|  | Bangladesh |  |  |  |  |
|  | Pakistan |  |  |  |  |
|  | Nepal |  |  |  |  |

**S2: DATA EXTRACTION**

 S2.1 Data extraction form

| **Assessment group** | **Questions** |
| --- | --- |
| **Availability** | Guideline Title |
|  | Authors |
|  | Issuing Organisation |
|  | Date of latest revision |
|  | Is the guideline adapted from another source? |
|  | Publication type (e.g., website, journal article, etc)  Intended audience? |
|  | Geographical aim (e.g., China, UK, USA)  Intended audience |
|  | What is the setting/income classification? (e.g., Low resource setting) |
| **Inclusivity** | What group of patients is the guideline aimed at? |
|  | Special populations included?  Is there specific guidance for pregnant patients?  Is there specific guidance for people living with HIV?  Is there specific guidance for children?  Is there specific guidance for older people?  Are all the above recommendations tied to clear evidence?  Any other details on population? |
| **Methodology** | How was the guideline developed? |
|  | Which key stakeholder groups involved?  How evidence based was each section?  Are there plans for the guideline to be reviewed? |
| **Scope** | Any PPE advice given?  Does the guideline recommend a specific therapeutic intervention (treatment or prophylaxis)?  Did the guideline provide supportive care recommendations?  Are routine investigations advised? |
|  | Is there clear advice on symptom management? e.g., analgesia including dose and duration |
|  | Any information regarding vaccination? |
|  | Are empirical antibiotics recommended? |
|  | Any specific therapeutic contraindications?  Advise tailored to disease severity? |
|  | Notes/other recommendations |
|  | Are all the above recommendations tied to clear evidence? |
|  | Other comments |

**S3: FIGURES and TABLES**

**Table S3.1:** Quality of the included clinical management guidelines

| Guideline | Country/Region | Year | Domain 1 (%) | Domain 2 (%) | Domain 3 (%) | Domain 4 (%) | Domain 5 (%) | Domain 6 (%) | Overall score |
| --- | --- | --- | --- | --- | --- | --- | --- | --- | --- |
| China (MoH) | China | 2003 | 8 | 8 | 6 | 22 | 17 | 0 | 1 |
| Dermatology Advisor | Global | 2017 | 19 | 19 | 16 | 56 | 15 | 33 | 1 |
| Dermnet | Global | 2014 | 58 | 17 | 16 | 61 | 0 | 8 | 2 |
| ECDC | Europe | 2019 | 6 | 33 | 20 | 53 | 19 | 8 | 1 |
| Emedicine | Global | 2020 | 42 | 31 | 6 | 47 | 10 | 0 | 1 |
| Ireland HPSC | Ireland | 2021 | 19 | 22 | 11 | 50 | 21 | 0 | 1 |
| Medscape | Global | 2019 | 17 | 44 | 32 | 64 | 8 | 46 | 2 |
| NCDC | Nigeria | 2019 | 86 | 3 | 8 | 86 | 13 | 13 | 3 |
| PHE(UKHSA) | England | 2019 | 28 | 19 | 7 | 44 | 13 | 0 | 1 |
| Singapore FETP | Singapore | 2020 | 67 | 47 | 20 | 61 | 29 | 0 | 3 |
| Taiwan CDC | Taiwan | 2009 | 19 | 50 | 15 | 44 | 10 | 0 | 1 |
| UpToDate | Global | 2021 | 42 | 47 | 84 | 67 | 8 | 67 | 6 |
| US CDC | USA | 2018 | 19 | 22 | 17 | 64 | 21 | 0 | 2 |
| WHO | Global | 2019 | 6 | 6 | 10 | 50 | 25 | 8 | 1 |
